# COVID-19 vaccination and Guillain-Barré syndrome: analyses using the National Immunoglobulin Database

**DOI:** 10.1101/2021.12.14.21267418

**Authors:** Ryan Y.S. Keh, Sophie Scanlon, Preeti Datta-Nemdharry, Katherine Donegan, Sally Cavanagh, Mark Foster, David Skelland, James Palmer, Pedro M. Machado, Stephen Keddie, Aisling S. Carr, Michael P. Lunn, BPNS/ABN COVID-19 Vaccine GBS Study Group

**Author notes:** Correspondence to: Michael P. Lunn, MRC Centre for Neuromuscular Diseases, National Hospital of Neurology and Neurosurgery, Queen Square, University College London Hospitals NHS Foundation Trust, London, UK.

## Abstract

Vaccination against viruses has rarely been associated with Guillain-Barré syndrome (GBS). An association with the COVID-19 vaccine is unknown. We performed a population-based study of National Health Service data in England and a multicentre surveillance study from UK hospitals, to investigate the relationship between COVID-19 vaccination and GBS.

Firstly, case dates of GBS identified retrospectively in the National Immunoglobulin Database from 8 December 2021 to 8 July 2021 were linked to receipt dates of a COVID-19 vaccines using data from the National Immunisation Management System in England. For the linked dataset, GBS cases temporally associated with vaccination within a 6-week risk window of any COVID-19 vaccine were identified. Secondly, we prospectively collected incident UK-wide (four nations) GBS cases from 1 January 2021 to 7 November 2021 in a separate UK multicentre surveillance database. For this multicentre UK-wide surveillance dataset, we explored phenotypes of reported GBS cases to identify features of COVID-19 vaccine-associated GBS.

996 GBS cases were recorded in the National Immunoglobulin Database from January to October 2021. A spike of GBS cases above the 2016-2020 average occurred in March-April 2021. 198 GBS cases occurred within 6 weeks of the first-dose COVID-19 vaccination in England (0.618 cases per 100,000 vaccinations, 176 ChAdOx1 nCoV-19 (AstraZeneca), 21 tozinameran (Pfizer), 1 mRNA-1273 (Moderna)). The 6-week excess of GBS (compared to the baseline rate of GBS cases 6-12 weeks after vaccination) occurs with a peak at 24 days post-vaccination; first-doses of ChAdOx1 nCoV-19 accounted for the excess. No excess was seen for second-dose vaccination. The absolute number of excess GBS cases from January-July 2021 was between 98-140 cases for first-dose ChAdOx1 nCoV-19 vaccination. First-dose tozinameran and second-dose of any vaccination showed no excess GBS risk. Detailed clinical data from 121 GBS patients were reported in the separate multicentre surveillance dataset during this timeframe. No phenotypic or demographic differences identified between vaccine-associated and non-vaccinated GBS cases occurring in the same timeframe.

Analysis of the linked NID/NIMS dataset suggests that first-dose ChAdOx1 nCoV-19 vaccination is associated with an excess GBS risk of 0.576 (95%CI 0.481-0.691) cases per 100,000 doses. However, examination of a multicentre surveillance dataset suggests that no specific clinical features, including facial weakness, are associated with vaccination-related GBS compared to non-vaccinated cases. The pathogenic cause of the ChAdOx1 nCoV-19 specific first dose link warrants further study.

## Introduction

The first year of the COVID-19 pandemic produced robust information on the neurological and neuropsychiatric sequelae of SARS-CoV-2 infection.^1^ In the peripheral nervous system, brachial neuritis, facial palsy and Guillain-Barré syndrome (GBS) were subjects of particular interest.

From January 2020, case reports and case series of patients with GBS occurring around the time of SARS-CoV-2 infection raised the possibility of a link between GBS and COVID-19.^2,3^ However, a large national study of the UK population as well as in Singapore showed a decreased incidence of GBS during the pandemic and failed to find a definitive link between GBS and COVID-19 infection.^4,5^

GBS became an adverse event of special interest (AESI) related to vaccination in the 1970s when an excess of GBS cases was detected in the United States during the 1976/1977 A/New Jersey/76 influenza (‘swine flu’) vaccination campaign within 6 weeks of vaccination.^6^ Serial epidemiological analyses established the rate of GBS attributable to the ‘swine flu’ vaccine was approximately 4.9-5.9 per million vaccines, mostly from 14–28 days post-vaccination.^6^ Although GBS has been identified in subsequent annual surveillance of influenza vaccination programmes at a rate of 1 to 1.6 per million doses,^7,8^ a pathogenic explanation has not been found. Expert consensus largely derived from the vaccination surveillance is that GBS risk attributable to vaccination exists for up to 6 weeks (42 days).^7^

The global rollout of COVID-19 vaccines triggered extensive monitoring, with GBS as an AESI. Very rare adverse events, not visible even in large clinical trials, can be identified when mass vaccination monitoring systems are in place, and particularly when a unique disease phenotype emerges. Vaccine-induced immune thrombotic thrombocytopenia (VIIT)^9^ and more recently myocarditis^10^ were identified this way, and are unique diseases. VIIT occurs most commonly in association with ChAdOx1 nCoV-19 (AstraZeneca) recombinant adenoviral vector vaccine. VIIT manifests as thromboses, including cerebral venous sinus thrombosis, with an estimated incidence of 1 case per 100,000 exposures.^9^ Myocarditis seems specific to the tozinameran (Pfizer) and mRNA-1273 (Moderna) vaccines.^10^

Non-replicating viral vectors can deliver vaccination antigen or other pharmaceuticals to the host. Adenoviruses are commonly used, and four current adenoviral vector COVID-19 vaccines are authorised in at least one country (AstraZeneca, Sputnik V, Janssen and Convidecia). Despite the single observational study of McNeil *et al*.^11^ suggesting a link of GBS to vaccination against adenovirus infection, adenovirus vectors are thought to be benign. ChAdOx1 nCoV-19 utilises a replication-deficient simian adenovirus vector designed to evade anti-human adenovirus neutralising antibodies to stimulate a robust immune response.

The UK COVID-19 vaccination programme began on the 8^th^ December 2020 with tozinameran, then ChAdOx1 nCoV-19 in January and subsequently mRNA-1273 vaccinations. Vaccination was delivered sequentially to cohorts of the most vulnerable and elderly followed by deciles of age. 50% of adults over 50 years of age had had their first vaccination by mid-February 2021.

We aimed to combine multiple national data sources and systematically investigate any temporal relationship between COVID-19 vaccination and excess cases of GBS during the UK COVID-19 vaccination programme. We retrospectively interrogated a large database of patients hospitalised with GBS in England, Scotland and Northern Ireland treated with immunoglobulin from the National Immunoglobulin Database (NID). Using the common NHS identifier, we combined the English data with data from the National Immunisation Management System (NIMS) on all COVID-19 vaccinations data in England. Separately, we characterised a large surveillance dataset of the incident UK GBS cases, presenting both after COVID-19 vaccination, and also without vaccination during the same period, recording the timing of onset after COVID-19 vaccination.

## Materials and methods

### Retrospective analyses of NID/NIMS datasets

All cases of GBS admitted to hospital in England, Scotland and Northern Ireland and considered for immunoglobulin treatment are recorded in the NID. Because NHS England (NHSE) procures the total immunoglobulin supply for the UK except Wales, and mandates that all immunoglobulin prescriptions are approved by the local clinical panel and reported onto the Immunoglobulin Database within 90 days of administration to facilitate repayment of immunoglobulin costs to the trusts, there is nearly 100% compliance with detailed recording of immunoglobulin use across the three UK regions.^12^ All cases are confirmed as GBS by the admitting clinician and reviewed and authorised by an independent Immunoglobulin Assessment Panel, although Brighton Criteria are not recorded.

We extracted NID GBS cases from 1 January to 31 October 2021 and recorded diagnosis, their unique identifier and date of first immunoglobulin prescription. These numbers were compared to the historical GBS cases recorded in the NID from 2016 to 2020.

UK Department of Health and Social Care (DHSC) guidance for GBS treatment recommends intravenous immunoglobulin (IVIg) as first line therapy for patients with Hughes Grade 4 or more (significant disability), disease progressing towards intubation or ventilation or with high probability of respiratory insufficiency (mEGRIS score ≥3) or predicted poor prognosis (mEGOS ≥4).^13^ Although plasma exchange (PLEX) is also considered a first line option, in practice, this is not readily available and is very seldom used. Utilising IVIg use as a proxy for GBS incidence under-estimates the true incidence of GBS, as milder cases are not treated. However, IVIg is estimated to be given to 86% of European and UK GBS cases,^14^ and the 2021 data can be compared to previous years with similar clinical behaviours for admitted patients.

The National Immunisation Management System (NIMS) database is a national point of care system for capturing vaccination data from England. Patients in the NIMS database are also registered with their NHS number. The COVID-19 vaccination data were linked to the English cases of GBS identified from the NID from 8 December 2020 to 8 July 2021 using the unique NHS identifier by NHS England. The start date was the commencement of the COVID-19 vaccination programme in the UK. Of note, the NIMS database captures information on vaccinations for England only (population: 55,980,000 prevalent persons in 2021); Scottish (5,470,000 prevalent persons) and Northern Irish (1,885,000 prevalent persons) GBS data were not included in this section of the analysis. At the time of analysis, data on immunoglobulin administration for GBS were available to mid-July 2021. To accommodate the need for a 6-week post-vaccination follow-up information on vaccinations up to and including 27 May 2021 was included. The exposed population, which was used to calculate GBS rates post-vaccination, was taken from weekly published cumulative counts of vaccine usage in England up to this date.

### Prospective surveillance study

We conducted a prospective surveillance study to compare the demographic and phenotypic characteristics of GBS cases reported from 1 January 2021 to 7 November 2021, comparing GBS cases reported as having received COVID-19 vaccination and cases without vaccination. We recognise the included cases are heavily influenced by reporting bias as there was significant interest in the possibility of vaccination-related GBS at the time. The surveillance study should not be used in direct comparison to the retrospective analyses of linked data described above.

Reports of GBS were submitted by members of the British Peripheral Nerve Society (BPNS) and the Association of British Neurologists (ABN), with regular reminders to collect information on hospital presentations of GBS during the study period. Data were entered into the International Neuromuscular COVID-19 database (https://www.ucl.ac.uk/centre-for-neuromuscular-diseases/news/2020/may/international-neuromuscular-covid-19-database), at the Centre for Neuromuscular Disease, where database questions had been adapted to allow for reporting of COVID-19 vaccine-related neuromuscular cases. Surveillance study data collection ended on 7 November 2021 to allow time for retrospective case reporting. Anonymised clinical data of demographics, GBS diagnostic criteria, vaccine details, and prior COVID-19 infection, symptoms and management were collected. Cases were classified according to Brighton Collaboration GBS Working Group criteria^15^ by the study team to describe level of diagnostic certainty were recorded as previously.^4^

Two cases were excluded from analysis as the reporting clinicians later informed us of a change in diagnosis.

### Statistical Analysis

Statistical analysis was performed using STATA 16^16^ and R v4.0.2/ v4.0.3 (R Core Team).^17^

An ecological analysis presenting the number of GBS cases identified in the NID by calendar month was used to compare the incidence of GBS in the UK across the years 2016-2020 with that seen in January-July 2021.

The Spearman’s rank correlation test was used to test for correlation between the time after the first vaccine dose and the incidence of GBS.

One-way ANOVA was used to compare yearly GBS rates within age deciles of the linked NID/NIMS data in 2021 compared to 2019 and 2020, to determine if age distribution of GBS differed compared to previous years, using ONS population estimates as the denominators for calculation of rates. As GBS numbers were only available until July 2021, an estimate of annual numbers was produced on the assumption of a stable GBS incidence through the year to enable comparison to prior years. ONS population estimates for 2021 were assumed to be the same as 2020.

For the prospective surveillance study, Chi-square and Kruskal-Wallis tests were used to test for correlations between patient demographics, GBS characteristics or treatment details, and exposure to COVID-19 vaccination within 6 weeks of GBS onset.

A significance value of *p*<0.05 was used throughout.

### Ethics

The UK Health Research Authority was consulted and advised that the study did not require review by an NHS Research Ethics Committee, as this was an analysis of previously collected, non-identifiable anonymised data.

### Data availability

Data are available on request to the corresponding author.

## Results

### Retrospective analyses of NID/NIMS datasets

#### Ecological analysis – GBS cases recorded in NID from 2016 to 2020

Between 2016 and 2020, the NID recorded a mean of 1283 immunoglobulin-treated cases of GBS per year (95%CI 1159-1408, mean 107 cases per month in the three participating UK nations). In England, a mean of 1148 GBS cases occurred annually between 2016 and 2020 (95%CI 1022-1274, or 3.1 GBS cases per day). These annualised case counts represent the vast majority of hospitalised GBS cases in England, Scotland and Northern Ireland, resulting in an estimated GBS incidence rate of 1.99 per 100,000 individuals per year (95%CI 1.79-2.18). This is comparable to previous European and North American studies with incidence rates of between 0.84-1.91 per 100,000 individuals per year.^18^ As previously described, the UK experienced an overall reduction in cases in 2020 during the height of the COVID-19 pandemic, resulting in the lowest case number (1053 cases in 2020) and estimated incidence (1.57 cases per 100,000 individuals per year) in the five-year period.^4^

Fig. 1 summarises monthly frequency of incident GBS cases in the England, Scotland and Northern Ireland from the years of 2016 to 2020, and compares these to the monthly incident case numbers from January to October 2021 during the UK vaccination programme. A total of 996 GBS cases were recorded from January to October 2021, fewer than observed from January-October 2016 to 2019 (pre-pandemic range: 1054-1182 cases) but higher than for January-October 2020 (pandemic range: 856 cases).

**Figure 1.**
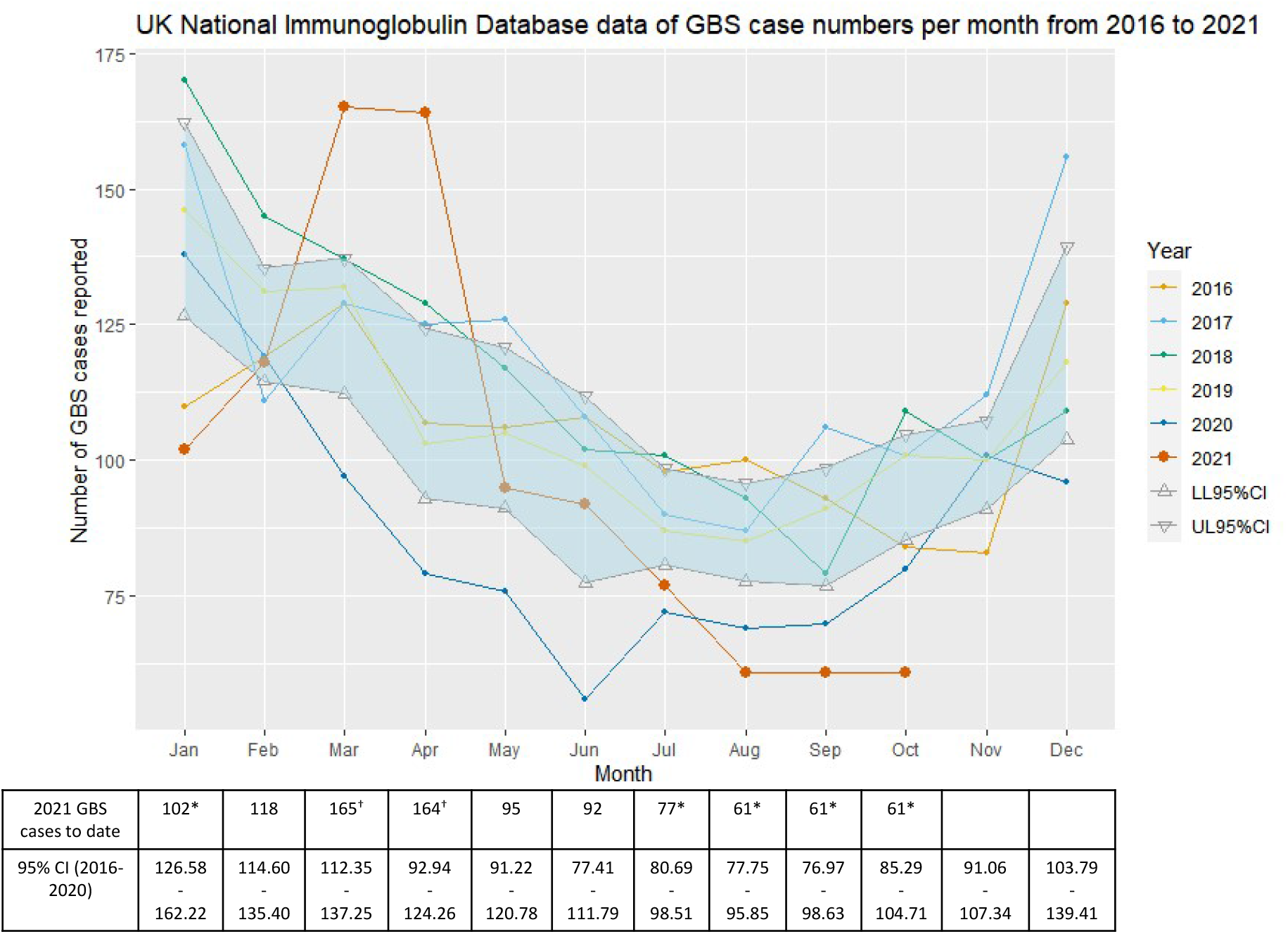
NHSE Immunoglobulin Database GBS cases 2016-2021. NHSE Immunoglobulin Database-derived numbers of incident GBS cases reported per month between 2016 and 2021 (year to date). The table below calculates mean monthly case numbers with 95% confidence intervals (CI) from 2016-2020 and compares them with absolute monthly case count for January-October 2021. *2021 GBS monthly number lower than 95% confidence interval of 2016 – 2020 monthly numbers; †2021 GBS monthly number higher than 95% confidence interval of 2016 – 2020 monthly numbers.

The number of GBS cases in January 2021 was significantly lower than the mean number seen in the same month 2016-2020, continuing the trend of lower GBS rates from 2020. However, England, Scotland and Northern Ireland experienced a spike of GBS cases in March and April 2021, before rates fell again below the lower range of the 95% CI for the 2016-2020 mean incident GBS case number in July to October 2021.

#### Analyses of linked NID/NIMS data

Annual GBS cases in England incident from 2019, 2020 and January to July 2021 were stratified by age decile, enabling age-specific incidence rate to be compared to recent years (Table 2). To enable a comparison across years, an estimate of annual case numbers for 2021 was produced based on assumption of minimal seasonal variation. An excess of cases in the 50-59 and 60-69 age groups was identified compared to 2019 and 2020, with statistically significant differences between age groups (*p*=0.00033).

**Table 1.**
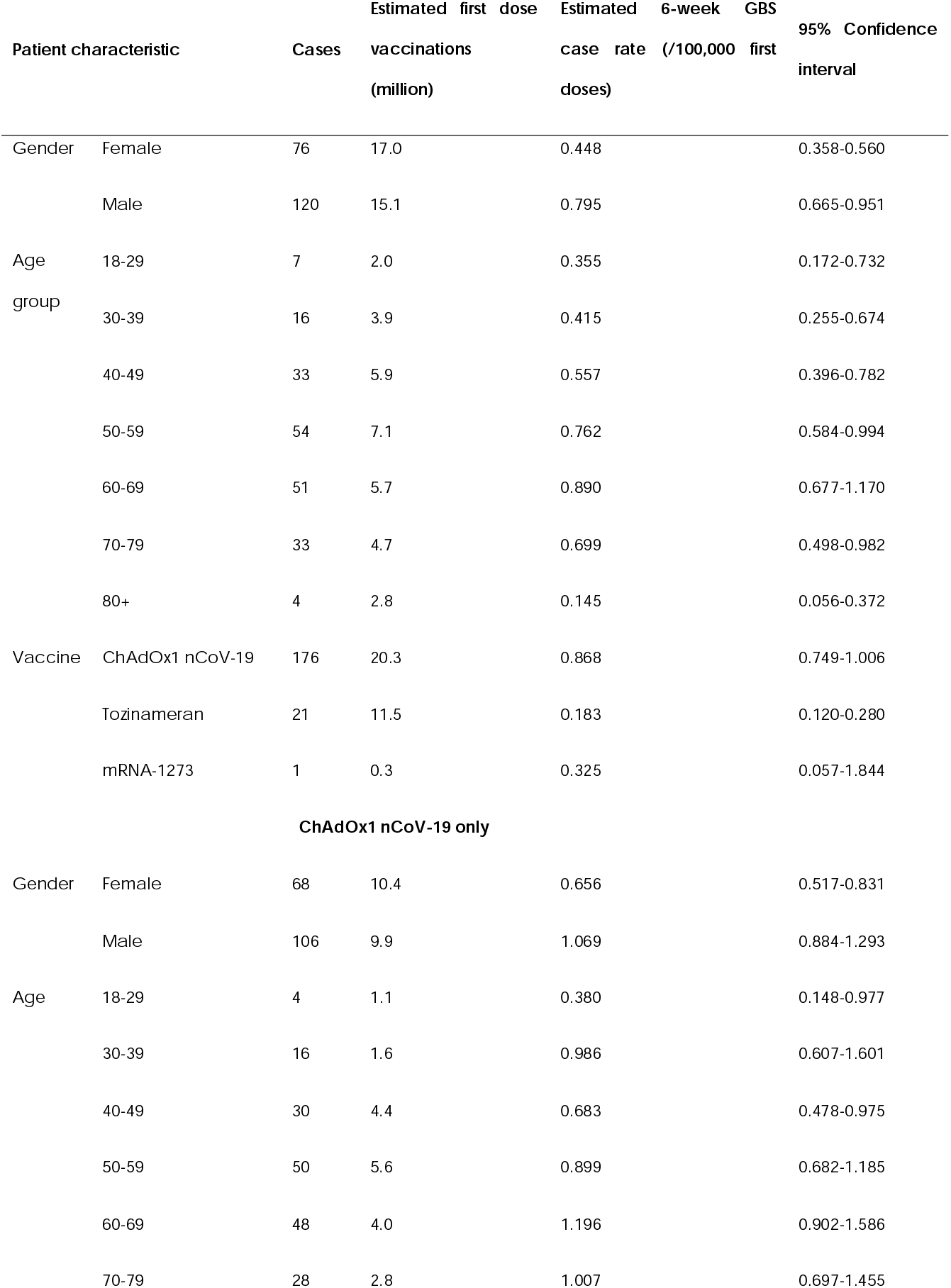
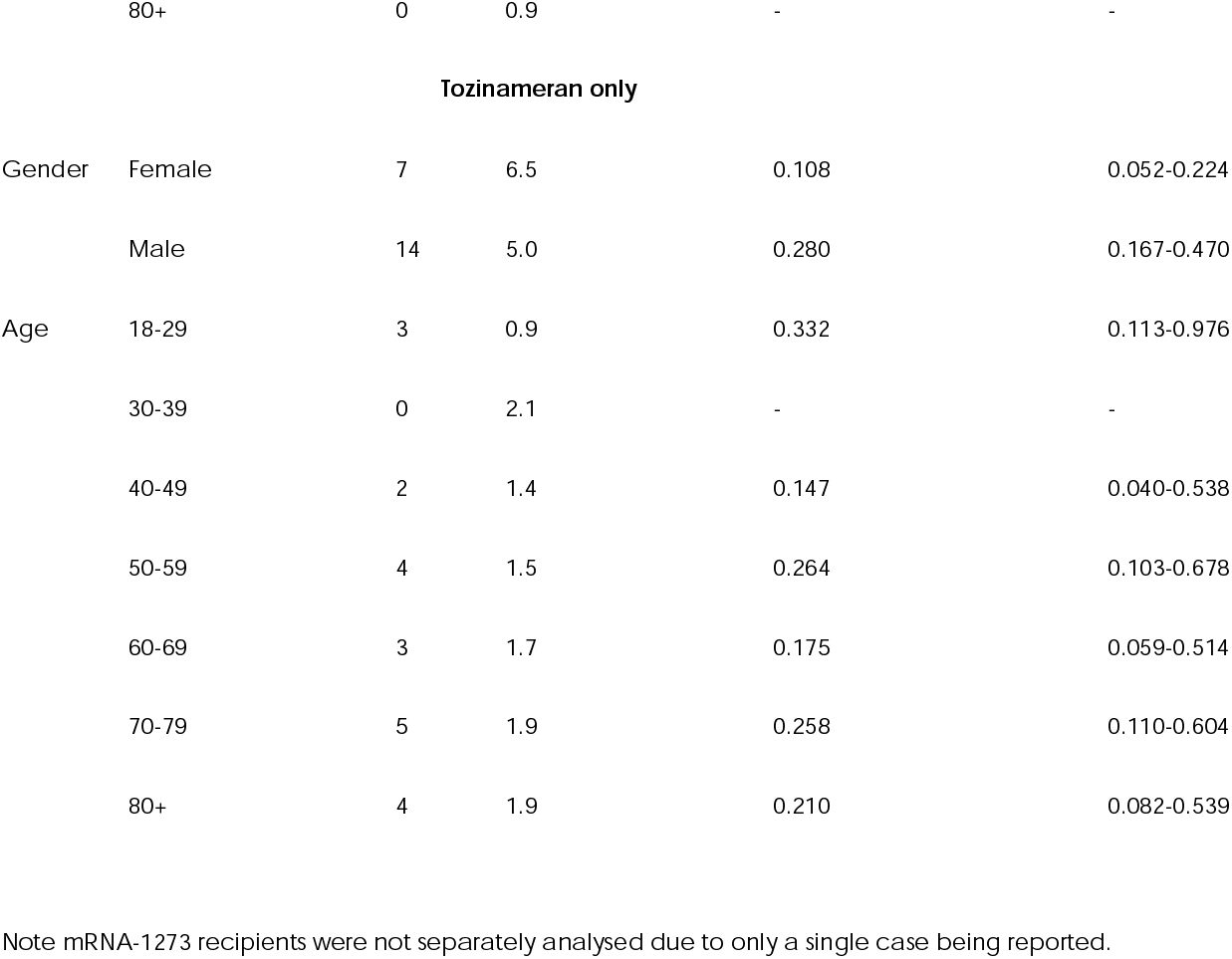
Patient characteristics of GBS cases documented in England occurring within 6 weeks of first dose COVID-19 vaccination with specific breakdown of gender and age for ChAdOx1 nCoV-19and Tozinameran recipients.

**Table 2.** GBS rates in England between 2019 and July 2021, separated by age group.

| Age group | Number of cases |  |  |  |  |  |  |  |  |
| --- | --- | --- | --- | --- | --- | --- | --- | --- | --- |
|  | 2019 |  |  | 2020 |  |  | 2021 January -July |  |  |
|  |  |  |  |  |  |  | *(Estimated annual 2021 total)* |  |  |
|  | GBS cases | ONS (million) | GBS rate per 100,000 patient years | GBS cases | ONS (million) | GBS rate per 100,000 patient years | GBS cases | ONS * (million) | GBS rate per 100,000 patient years |
| 18-29 | 109 | 8.55 | 1.28 | 90 | 8.48 | 1.06 | 38 (65) | 8.48 | 0.77 |
| 30-39 | 117 | 7.54 | 1.55 | 99 | 7.56 | 1.31 | 48 (82) | 7.56 | 1.09 |
| 40-49 | 120 | 7.13 | 1.68 | 110 | 7.11 | 1.55 | 84 (144) | 7.11 | 2.04 |
| 50-59 | 191 | 7.58 | 2.52 | 138 | 7.64 | 1.81 | 161 (276) | 7.64 | 3.63 |
| 60-69 | 213 | 5.91 | 3.60 | 200 | 5.98 | 3.34 | 145 (248) | 5.98 | 4.18 |
| 70-79 | 202 | 4.72 | 4.28 | 163 | 4.82 | 3.38 | 101 (173) | 4.82 | 3.61 |
| 80+ | 98 | 2.84 | 3.45 | 60 | 2.86 | 2.10 | 27 (46) | 2.86 | 1.63 |

Estimated annual total for 2021 (*italics*) is extrapolated assuming stable GBS incidence across the year based on numbers from January to July 2021. \*For 2021 ONS population estimates were assumed to be the same as 2020. ONS = Office of National Statistics population estimates; GBS rate = incidence rate of GBS cases (per 100,000 patient years, assuming that each individual in the ONS population estimate is followed for a full year, or in the case of 2021 estimates for 7 months)

Using the linked NID/NIMS data, the first record of GBS occurring within 6 weeks after a COVID-19 vaccination was in January 2021. Of note, not all GBS patients in NID had a vaccination record. 198 cases of GBS identified in the linked NID/NIMS data study period occurred within 6 weeks of the first dose of any COVID-19 vaccine (0.618 cases per 100,000 vaccinations in 6 weeks, all ages). A total of 32.1 million first dose vaccinations were recorded during the reporting period (20.3 million ChAdOx1 nCoV-19, 11.5 million tozinameran and 0.3 million mRNA-1273). Of the 198 linked GBS cases, 176 followed a first dose ChAdOx1 nCoV-19 vaccine (rate 0.868 per 100,000) and 21 followed a first dose tozinameran vaccine (rate 0.183 per 100,000). Only one case was reported within 6 weeks of mRNA-1273vaccination. Only 23 GBS cases were reported within 6 weeks of any second vaccine dose.

Table 1 and Fig. 2 summarise patient characteristics of GBS cases in England occurring within 6 weeks of first COVID-19 vaccination. The GBS incidence after first vaccination was highest in males receiving the ChAdOx1 nCoV-19vaccine (1.069 per 100,000 doses).

**Figure 2.**
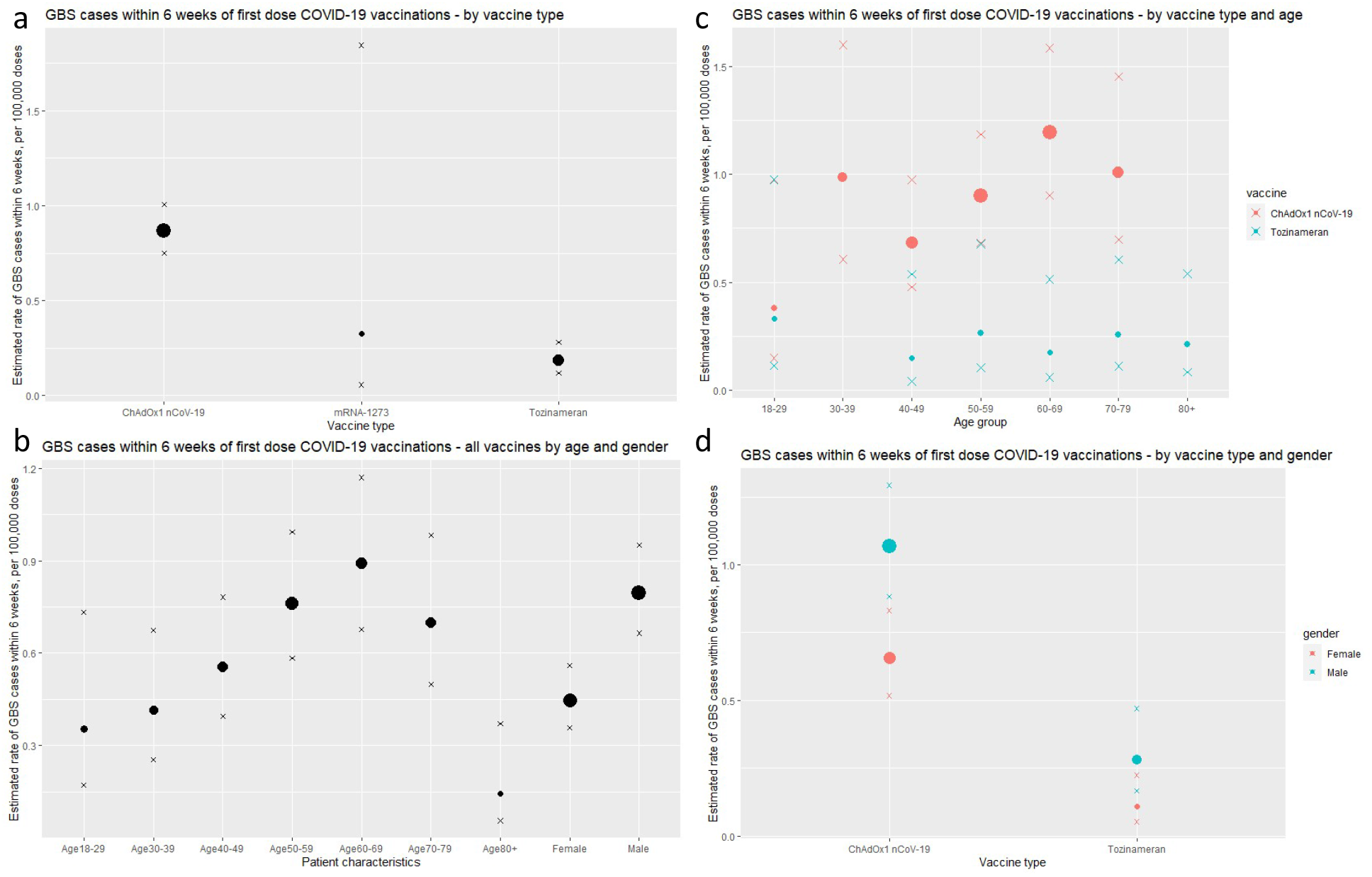
GBS case rate within 6 weeks of first-dose COVID-19 vaccination in England. Estimated rate of GBS cases within 6 weeks of first COVID-19 vaccination (per 100,000 doses), separated (a) by vaccine type, (b) by age/gender, (c) by age for ChAdOx1 nCoV-19 and tozinameran vaccines, and (d) by gender for ChAdOx1 nCoV-19and tozinameran vaccines. Size of dots weighted based on GBS case numbers. Crosses represent upper and lower limits of 95% confidence intervals.

The daily number of incident GBS cases, with an 84-day post-vaccine follow-up from dose 1 and 2 of COVID-19 vaccination was plotted (Fig. 3). A peak of GBS cases is observed around 24 days following a first dose, with higher numbers of cases seen in the period of 2 to 4 weeks after vaccination than in other periods. First doses of ChAdOx1 nCoV-19vaccine account for the majority of this increase. A similar pattern is not seen following a second dose of any vaccine. Using the Spearman’s rank correlation test of randomness, the occurrence of GBS was random for all times after the first-dose tozinameran vaccine (*p*=0.84) (no peak associated with tozinameran vaccine) and after the second-dose vaccination of all vaccines (*p*=0.85). However, it was non-random for ‘all first-dose’ vaccination (*p*=0.009) and the first dose of the ChAdOx1 nCoV-19 vaccination (*p*=0.004).

**Figure 3.**
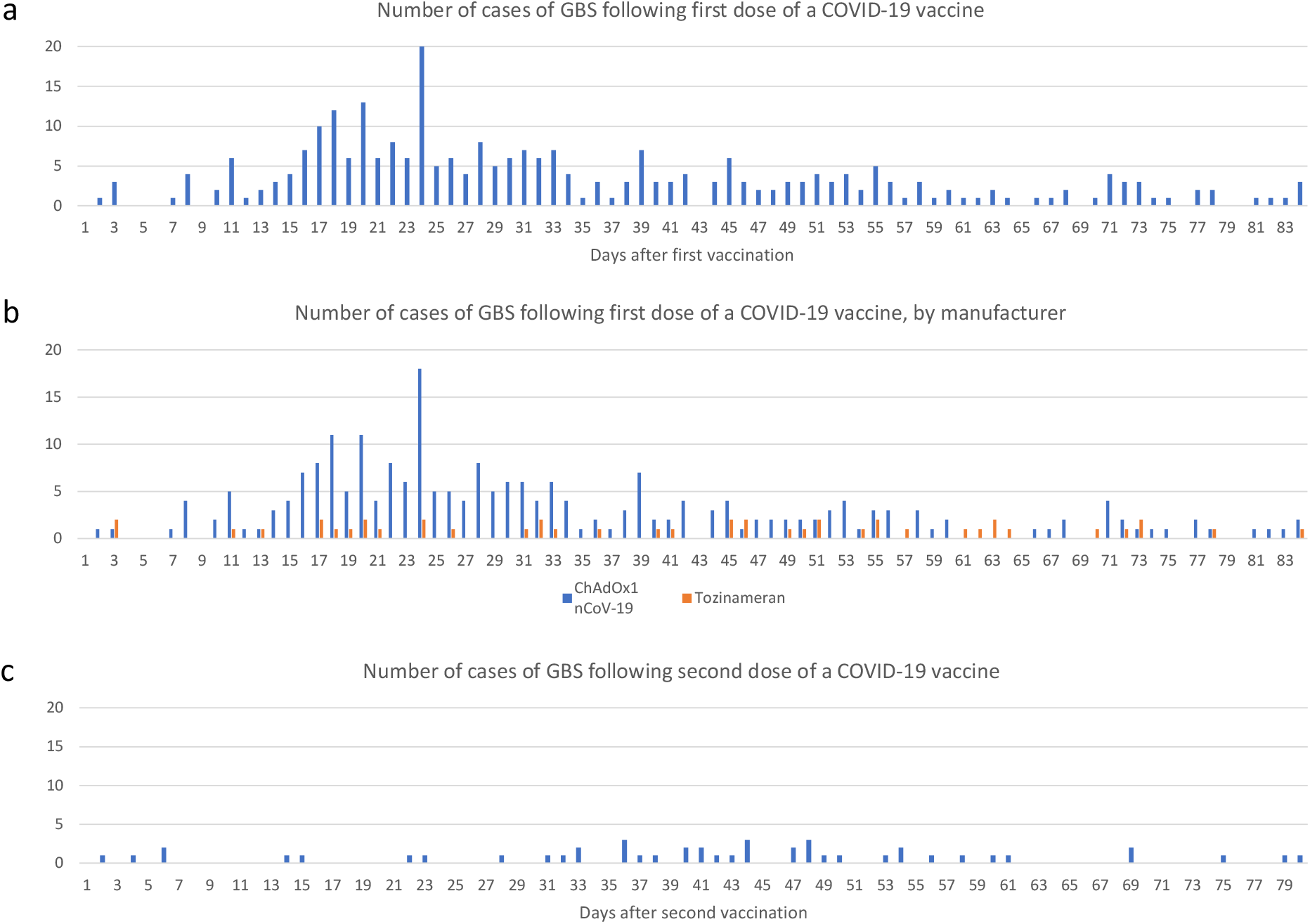
GBS case numbers in England by day following vaccination. Number of GBS cases 0 to 84 days following (a) any first dose vaccination, (b) first dose stratified by vaccine brand and (c) second dose COVID-19 vaccination.

Using case numbers from day 43-84 after first-dose vaccination as a comparison group (assuming this group represents a baseline random GBS rate), the excess risk in the first 42 days post-ChAdOx1 nCoV-19 vaccine is 0.576 GBS cases per 100,000 doses (*95% CI* 0.481-0.691). With an estimated 20.3 million first ChAdOx1 nCoV-19 doses given at the time of analysis, this suggests an absolute number of 98-140 excess GBS cases. There was no significant difference between the GBS cases associated with the first dose of tozinameran and second-dose vaccination numbers in the day 0-42 or day 43-84 case numbers. Fig. 4 summarises the estimated excess risk for different vaccinated groups.

**Figure 4.**
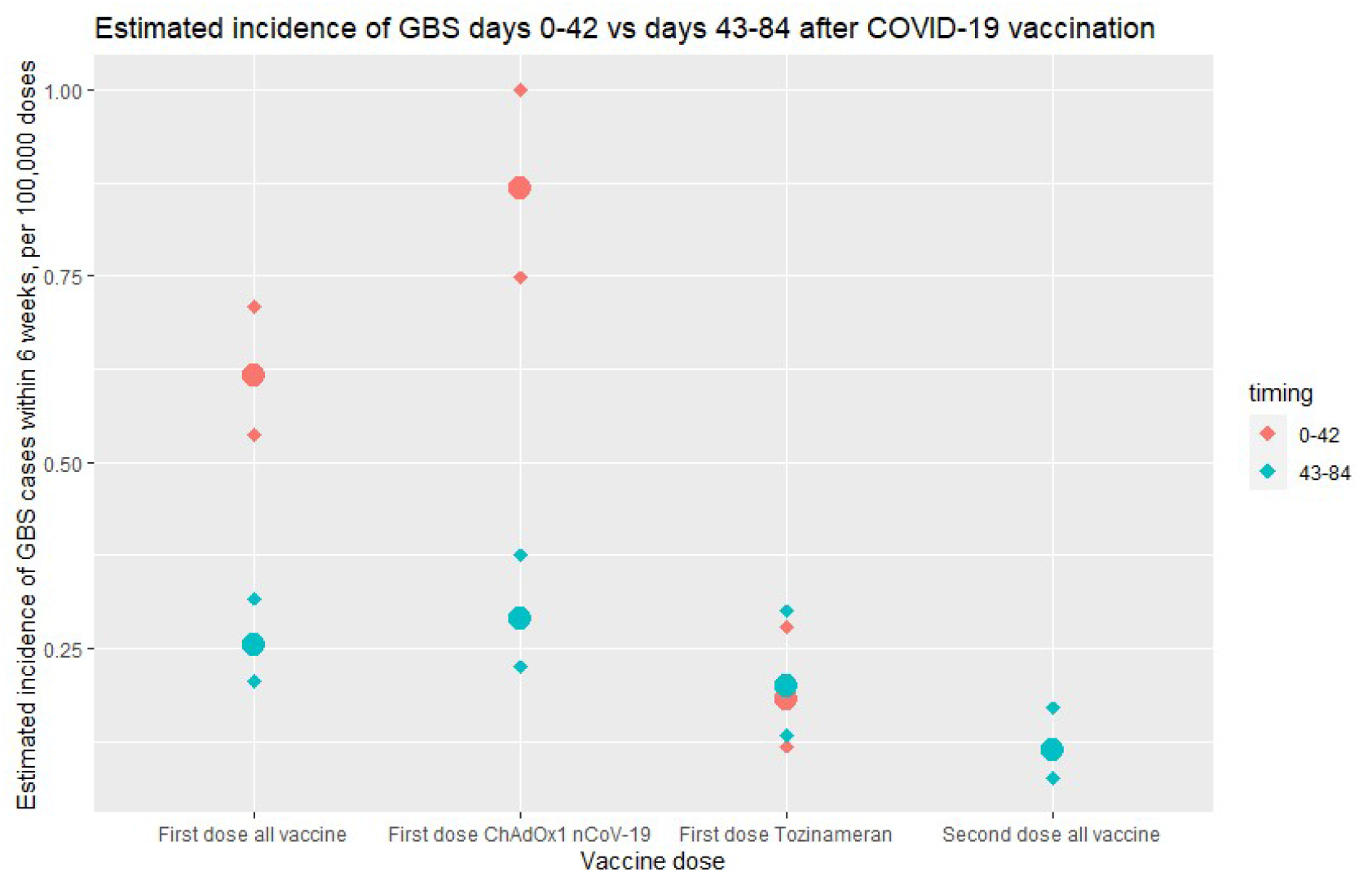
Excess risk in first 42 days following vaccination in England. Estimated incidence of GBS cases within 6 weeks (per 100,000 vaccine doses), comparing reports of GBS cases 0-42 days (red) and 43-84 (blue) for first-dose vaccines (all vaccines, ChAdOx1 nCoV-19, tozinameran) and second-dose vaccines. Diamonds represent upper and lower limits of 95% confidence intervals. An excess of GBS cases is noted in the first 42 days following first-dose vaccination, accounted for by the ChAdOx1 nCoV-19vaccine.

### Prospective surveillance study

Between 1 January and 7 November 2021, 121 cases of GBS were reported by the BPNS/ABN network. Since clinicians and the public were highly sensitised to vaccination and concerns about risk, the reporting of this dataset is biased in favour of vaccine-related cases. The median age of reported cases was 59 years, with 59% being male. 106 patients (87.3%) had received COVID-19 vaccination prior to GBS onset, with 80 (66.1% of the total dataset) having received a first-dose vaccination within 42 days of GBS onset. In comparison, from the linked NID/NIMS dataset, 198 of the 659 GBS cases in England during this timeframe (30%) were reported to be within 42 days of vaccination. This highlights the reporting bias of the dataset towards vaccine-associated GBS. As expected with the timing of the vaccine rollout, 90% of GBS cases reported from January to April 2021 were within 6 weeks of vaccination, compared to only 35% of cases from May 2021 onwards.

Further demographic and clinical characteristics of the patients described within the dataset are shown in Table 3.

**Table 3.**
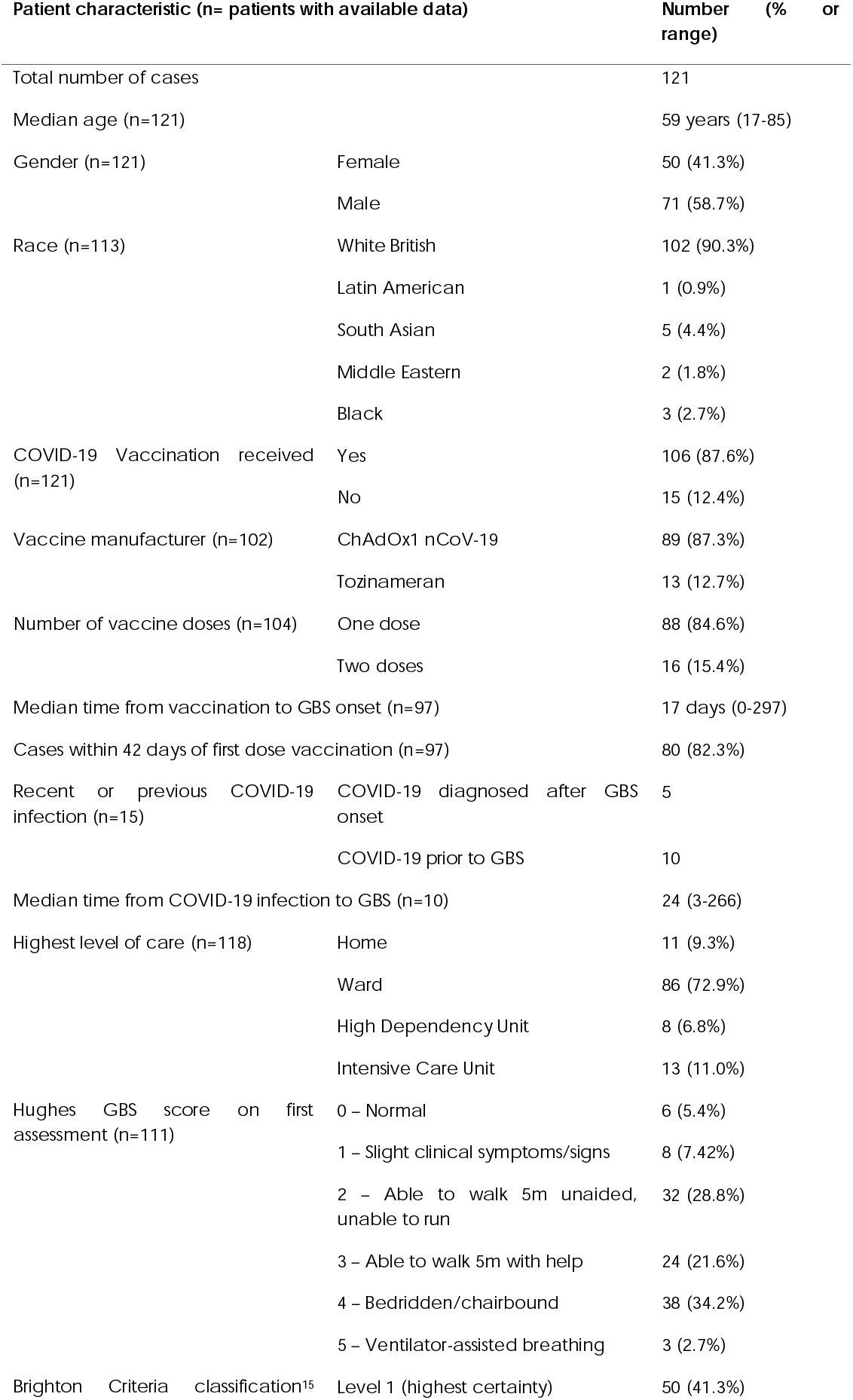
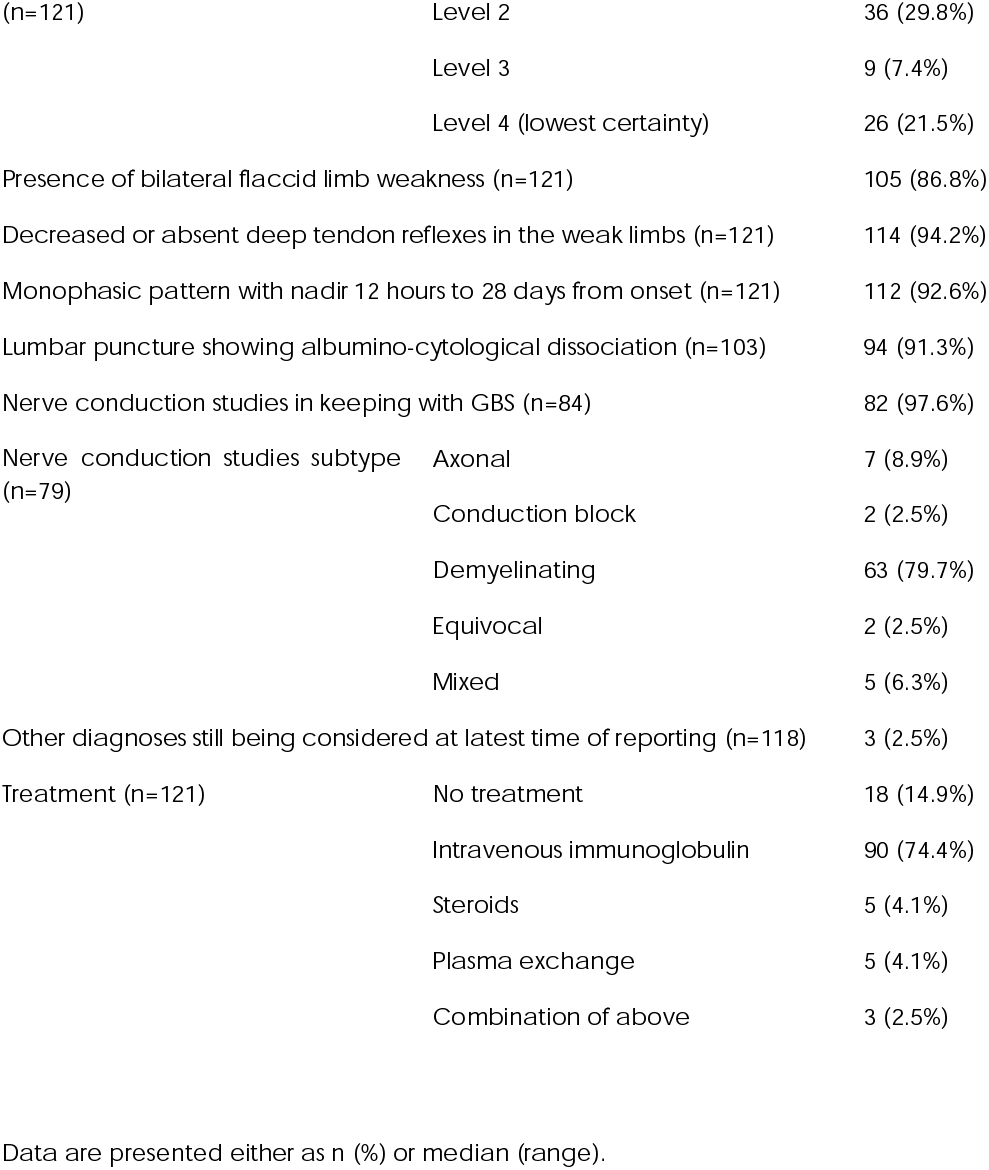
Characteristics of patients from clinician-reported surveillance study.

| Patient characteristic (n= patients with available data) |  | Number | (% or range) |
| --- | --- | --- | --- |
| Total number of cases |  | 121 |  |
| Median age (n=121) |  | 59 years | (17-85) |
| Gender (n=121) | Female | 50 | (41.3%) |
|  | Male | 71 | (58.7%) |
| Race (n=113) | White British | 102 | (90.3%) |
|  | Latin American | 1 | (0.9%) |
|  | South Asian | 5 | (4.4%) |
|  | Middle Eastern | 2 | (1.8%) |
|  | Black | 3 | (2.7%) |
| COVID-19 Vaccination received (n=121) | Yes | 106 | (87.6%) |
|  | No | 15 | (12.4%) |
| Vaccine manufacturer (n=102) | ChAdOx1 nCoV-19 | 89 | (87.3%) |
|  | Tozinameran | 13 | (12.7%) |
| Number of vaccine doses (n=104) | One dose | 88 | (84.6%) |
|  | Two doses | 16 | (15.4%) |
| Median time from vaccination to GBS onset (n=97) |  | 17 days | (0-297) |
| Cases within 42 days of first dose vaccination (n=97) |  | 80 | (82.3%) |
| Recent or previous COVID-19 infection (n=15) | COVID-19 diagnosed after GBS onset | 5 |  |
|  | COVID-19 prior to GBS | 10 |  |
| Median time from COVID-19 infection to GBS (n=10) |  | 24 | (3-266) |
| Highest level of care (n=118) | Home | 11 | (9.3%) |
|  | Ward | 86 | (72.9%) |
|  | High Dependency Unit | 8 | (6.8%) |
|  | Intensive Care Unit | 13 | (11.0%) |
| Hughes GBS score on first assessment (n=111) | 0 – Normal | 6 | (5.4%) |
|  | 1 – Slight clinical symptoms/signs | 8 | (7.42%) |
|  | 2 – Able to walk 5m unaided, unable to run | 32 | (28.8%) |
|  | 3 – Able to walk 5m with help | 24 | (21.6%) |
|  | 4 – Bedridden/chairbound | 38 | (34.2%) |
|  | 5 – Ventilator-assisted breathing | 3 | (2.7%) |
| Brighton Criteria classification<sup>15</sup> | Level 1 (highest certainty) | 50 | (41.3%) |

|  |  |  |
| --- | --- | --- |
| (n=121) | Level 2 | 36 (29.8%) |
|  | Level 3 | 9 (7.4%) |
|  | Level 4 (lowest certainty) | 26 (21.5%) |
| Presence of bilateral flaccid limb weakness (n=121) |  | 105 (86.8%) |
| Decreased or absent deep tendon reflexes in the weak limbs (n=121) |  | 114 (94.2%) |
| Monophasic pattern with nadir 12 hours to 28 days from onset (n=121) |  | 112 (92.6%) |
| Lumbar puncture showing albumino-cytological dissociation (n=103) |  | 94 (91.3%) |
| Nerve conduction studies in keeping with GBS (n=84) |  | 82 (97.6%) |
| Nerve conduction studies subtype (n=79) | Axonal | 7 (8.9%) |
|  | Conduction block | 2 (2.5%) |
|  | Demyelinating | 63 (79.7%) |
|  | Equivocal | 2 (2.5%) |
|  | Mixed | 5 (6.3%) |
| Other diagnoses still being considered at latest time of reporting (n=118) |  | 3 (2.5%) |
| Treatment (n=121) | No treatment | 18 (14.9%) |
|  | Intravenous immunoglobulin | 90 (74.4%) |
|  | Steroids | 5 (4.1%) |
|  | Plasma exchange | 5 (4.1%) |
|  | Combination of above | 3 (2.5%) |

Data are presented either as n (%) or median (range).

Forty-two patients (34.7%) were reported to have facial weakness in association with other GBS findings. Facial weakness was bilateral in 37 of these patients. Only 7 patients (5.8%) had pure bilateral facial paralysis with paraesthesia.

Only four patients were reported with positive ganglioside antibodies (GM1, GM2, GM1/GD1b, GD1a/GD1b), but this was likely to have been under-reported because of the time delays in anti-ganglioside results being available and inconsistent testing. Equally, limited COVID access to neurophysiology services may explain why only 84 of the 121 patients had nerve conduction studies; these data deficiencies limit the Brighton Collaboration diagnostic categorisation.

Comparing surveillance study patients who had first-dose vaccination within 42 days of GBS onset to unvaccinated patients and patients who had GBS more than 42 days after first-dose vaccination, no significant differences were found in terms of gender (*p*=0.54), age (*p*=0.25), highest level of care (*p*=0.36), Hughes GBS disability score on admission (*p*=0.75), Brighton level of diagnostic certainty (*p*=0.53), nerve conduction study changes (*p*=0.44), treatment (*p*=0.52) or presence of unilateral or bilateral facial weakness *(p*=0.77).

## Discussion

We have presented parallel studies designed to investigate a possible relationship between COVID-19 vaccination and GBS. The first study linked nationalised databases and the second was based on prospective case reporting.

These data suggest a clear and plausible excess of GBS cases occurring within 42 days after the first dose of ChAdOx1 nCoV-19 COVID-19 vaccination. The complexities of timing and delivery of multiple vaccines to age and at-risk cohorts of patients in the UK, on a naturally unstable GBS baseline make accurate identification and quantification of the risks difficult in real time. The data provide an estimate of 5.8 cases of GBS (4.8 – 6.9) per million first doses of ChAdOx1 nCoV-19 vaccine and no measurable excess of GBS associated with first doses of tozinameran. This equates to an absolute excess of between 98 and 140 cases of GBS attributable to ChAdOx1 nCoV-19 vaccination up to 8 July 2021 in England. This should be compared to estimates that the vaccination programme has directly averted over 52,600 hospitalisations, between 21.3 and 22.9 million infections and between 57,500 and 62,700 deaths over the same time period.^19^

We decided to use GBS case numbers from day 43–84 after vaccination as a comparator group, rather than an externally-derived control based on historical GBS numbers, as GBS case numbers during the COVID-19 pandemic may be different from historical baselines.^4^ GBS incidence in this group was estimated at 1.9 cases per day, lower than the historical 2016-2020 average of 3.1 cases per day based on NID data in England, supporting the hypothesis that the baseline GBS case numbers in 2021 continue to be lower than pre-pandemic levels.

The total number of cases of GBS in the NID from January to October 2021 is lower than the 10-month total of January to October 2016 to 2019; but is higher than total case numbers during 2020. A monthly increase in GBS cases in March and April 2021 was notable, but total numbers fell back into the normal range thereafter and thus this ‘spike’ is the only hint of a causative link in simple occurrence data.

The spike in case numbers and the subsequent return to normal levels might be explained in several ways. The baseline onto which any vaccination-related cases are built varies because of the normal seasonal reduction of GBS, but also the unknown effects of ongoing social distancing measures, which reduced the 2020 cases of GBS by about 1/3. Social distancing and other lockdown measures in the UK were slowly relaxed in the first half of 2021 and some GBS increase might be expected through greater pathogen exposure. In addition, vaccinated individuals may have rapidly changed their social behaviour after vaccinations, increasing exposure to other infections which are known to increase GBS risk, but this behaviour would have been expected to continue and grow as more were vaccinated, which is not the pattern demonstrated here. The remaining nationally-mandated social distancing measures may continue to account for the lower than usual numbers of GBS cases in July – October 2021.

If all COVID-19 vaccines were associated with an increased risk of GBS across all age groups, a more sustained rise in GBS incidence would be expected. 9,646,715 people, mostly the older and more vulnerable, were vaccinated from early December 2020 to 1^st^ February, but the increase in GBS cases was not seen until mid-February.

ChAdOx1 nCoV-19 vaccinations commenced in early January, quickly overtaking tozinameran as the predominant vaccine administered, to a significant proportion of the apparently more susceptible 50- to 70-year-old age group, with 24,858,665 adults receiving the first dose of COVID-19 vaccine from 1 Feb 2021 to 1 May 2021 (representing 50,5% of the UK population).^20^

Interestingly, there is no recognisable increase in GBS after the second dose which may be a pathophysiological phenomenon, individual susceptibility, or because patients experiencing adverse effects from the first dose did not take the second vaccine dose. We know of a single patient who was reported to have GBS-like illness following both first- and second-dose of the ChAdOx1 nCov-19 vaccine. The patient initially developed a facial diplegia and paraesthesia phenotype with subsequent gait disturbance and elevated CSF protein, and improved with IVIg. Two months later, two weeks after their second dose of ChAdOx1 nCoV-19 vaccination, they developed increasing weakness with neuropathic pain, with elevated CSF protein, demyelinating changes on nerve conduction studies and MRI enhancement of the cauda equina, with only partial response to IVIg treatment.

The single patient in our surveillance study who had recurring neuropathic symptoms after second-dose vaccination appears to be a unique report in the UK at this time, and may potentially represent acute-onset chronic inflammatory demyelinating polyradiculoneuropathy (A-CIDP) rather than true GBS. A recent study from Israel of COVID-19 vaccinations in patients with previous GBS described a single patient with recurrent GBS-like illness time-linked to both doses of their tozinameran vaccination, but limited information is available regarding the strength of GBS diagnosis.^21^

A small excess of GBS in males is seen in this vaccine-associated GBS cohort; as is documented in the GBS literature,^15^ but the reason for this remains unclear. Furthermore, the phenotype of the reported cases of GBS in the surveillance study gives no recognisable vaccine-specific GBS features unlike the Vaccine-Induced Immune Thrombotic Thrombocytopenia (VITT), with distinguishable clinical features and a biomarker in the anti-PF4 antibodies.^11^ Cases of severe (bi)-facial palsy have been widely discussed,^22-25^ yet this feature does not occur more often than in the non-vaccine associated cases in our dataset.

The reason for the association between only ChAdOx1 nCoV-19 vaccination and GBS is unclear. COVID-19 infection does not have a strong, or possibly any, increased risk of GBS,^4,5^ and the lack of increased risk associated with tozinameran vaccination implies that it is unlikely that the COVID-19 spike protein is the causative factor for the increased risk. The excess incidence is estimated to be 5.8 cases per million doses, similar to the estimates for the 1976 ‘swine flu’ vaccine and higher (but within the same order of magnitude) as the reported excess cases for the modern influenza and yellow fever vaccines. It is far below the 1:1000 cases of GBS in *C. jejuni* gastroenteritis or Zika-virus. A non-specific immune activation in susceptible individuals might therefore be implicated, but if that were the case similar risks might apply to all vaccine types. It is therefore logical to suggest the simian adenovirus vector may account for the increased risk. Adenoviruses have not been strongly associated with GBS in previous studies,^26^ and any association between adenoviral vaccination and GBS^11^ has only once been reported. Nevertheless, adenovirus testing is not routinely performed in cases of GBS in the UK, and whether they may account for a proportion of ‘idiopathic’ or ‘serology negative’ GBS may be the subject for further study.

Although the first retrospective analysis presented here employs two cross-referenced national and mandated datasets, there are still confounders, bias and criticisms as applicable to many epidemiological studies. We were unable to individually validate cases of immunoglobulin-treated GBS. However, these patients would need to be unwell enough to be admitted to hospital, and the diagnosis was assessed not only by the admitting physicians but also by an Immunoglobulin Assessment Panel who authorise the treatment. Furthermore, the data are controlled against data from earlier years where GBS rates are consistent with other international, methodologically robust epidemiological studies with high levels of ascertainment and clinical validity. We recognise that patients with ‘mild GBS’ may not attend hospital or be treated, but this has been the same in past years. We also recognise that patients may have been more reluctant to attend hospital, although this was not obviously observed in 2021, and paralysed patients are likely to have attended more than those with lesser disability.

We explored detailed phenotypes by collecting data on GBS presentations to UK neurologists with a continuation of the BPNS/ABN surveillance study. 121 cases were reported, representing only 13% of GBS cases in that period of 2021. Clinicians were much more likely to report cases with recent vaccination compared to those who were unvaccinated. Nonetheless, this dataset allowed for deeper confirmation of GBS diagnoses, with 79% of the cohort meeting Brighton Collaboration diagnostic criteria level 1, 2, or 3 (compared to 15% of the cases reported to the MHRA ‘Yellow Card’ system [personal communication]). Within our dataset, there were no differences identified between patients who had recent vaccination and those who had not, in terms of baseline demographics, disease course or treatment. Although BPNS and ABN members reported cases of facial weakness associated with GBS, there was no increase linked to vaccination status. The description of some of these cases best fit the ‘atypical’ GBS presentation of ‘bilateral facial weakness with paraesthesia’, which is felt to represent <5% of total GBS cases.^27^

Another recently published UK-based study has analysed neurological complications in the context of recent COVID-19 vaccination and infection, and has reported comparable excess in GBS cases of 3.8 per million doses ChAdOx1 nCoV-19 vaccine. The incidence rate ratio (IRR) of 2.90 at 15-21 days post-vaccination suggests a similarly plausible time-locked association to that which we observed, in keeping with the pathological mechanism of GBS.^28^ However, they also report a possible increase in GBS cases in relation to COVID-19 infection of 14.5 per million COVID-19 infections, with an IRR of 5.25.^28^ We note that neurological manifestations were identified using hospital coding data, which may be less accurate than NID identification of Immunoglobulin Panel-scrutinised GBS cases. Moreover, mortality rate was 1.8% in that cohort, lower than the 3-10% mortality generally associated with GBS^27^. In addition, 16% of COVID-19-associated GBS in their cohort (7/43 cases) were diagnosed on the same day as COVID-19 diagnosis, which makes it less likely that GBS was caused by a post-infectious immune process triggered by COVID-19 infection. For these and other reasons, we believe that our study’s findings on excess risk may provide a more accurate estimate of risk.

Our study reports an association between first-dose ChAdOx1 nCoV-19COVID-19 vaccination and GBS, accounting for an estimated excess incidence of 5.8 GBS cases per million first doses. The cause for this association remains unclear, and excess risk remains comparable to previous vaccine-associated GBS. The risk in proportion to the benefits of vaccination is very small. Further studies are required to confirm these observations, to determine causality, to explore the pathogenic mechanisms and to investigate effects of other COVID-19 vaccine preparations in use elsewhere in the world.

## Data Availability

Data are available on request to the corresponding author.

## Abbreviations

ABN: Association of British Neurologists
AESI: Adverse Event of Special Interest
BPNS: British Peripheral Nerve Society
DHSC: Department of Health and Social Care
GBS: Guillain-Barré syndrome
NHS: National Health Service
NHSE: National Health Service England
NID: National Immunoglobulin Database
NIMS: National Immunisation Management System
VIIT: Vaccine-induced immune thrombotic thrombocytopenia

## Acknowledgements

This section is not mandatory.

### Appendix 1

BPNS/ABN COVID-19 Vaccine GBS Study Group:

Hadi Manji, National Hospital of Neurology and Neurosurgery, Queen Square, University College London Hospitals NHS Foundation Trust, London

Tim Lavin, Manchester Centre for Clinical Neurosciences, Salford Royal NHS Foundation Trust, Manchester

James B. Lilleker, Manchester Centre for Clinical Neurosciences, Salford Royal NHS Foundation Trust, Manchester

David Gosal, Manchester Centre for Clinical Neurosciences, Salford Royal NHS Foundation Trust, Manchester

Robert D.M. Hadden, Kings College Hospital NHS Foundation Trust, London

Taylor Watson-Fargie, Institute of Neurological Sciences, Queen Elizabeth University Hospital, NHS Greater Glasgow and Clyde, Glasgow.

Kathryn Brennan, Institute of Neurological Sciences, Queen Elizabeth University Hospital, NHS Greater Glasgow and Clyde, Glasgow.

Andreas Themistocleous, Nuffield Department of Clinical Neurosciences, University of Oxford, Oxford, UK

Jacquie Deeb, Queens Hospital, Barking, Havering and Redbridge University Hospitals NHS Trust, Romford

Ana Romeiro, Wexham Park Hospital, NHS Frimley Health Foundation Trust, Slough

Puja Mehta, Kings College Hospital NHS Foundation Trust, London; UCL Queen Square Institute of Neurology, London

Dimitri Kullmann, National Hospital for Neurology and Neurosurgery, Queen Square, University College London Hospitals NHS Foundation Trust, London

James Miller, Royal Victoria Infirmary, The Newcastle upon Tyne Hospitals NHS Foundation Trust, Newcastle upon Tyne

Amar Elsaddig, Manchester Centre for Clinical Neurosciences, Salford Royal NHS Foundation Trust, Manchester

Adam Molyneux, John Radcliffe Hospital, Oxford University Hospitals NHS Foundation Trust, Oxford

Plamen Georgiev, Broomfield Hospital, Mid and South Essex NHS Foundation Trust, Chelmsford

Aaron Ben-Joseph, Maidstone Hospital, Maidstone and Tunbridge Wells NHS Trust, Kent

James Holt, The Walton Centre NHS Foundation Trust, Liverpool

Jacob Roelofs, Royal Victoria Infirmary, The Newcastle upon Tyne Hospitals NHS Foundation Trust, Newcastle upon Tyne

Fadi Alkufri, Kent and Canterbury Hospital, East Kent Hospitals University NHS Foundation Trust

David Allen, Southampton General Hospital, University Hospital Southampton NHS Foundation Trust, Southampton

Simon Shields, Somerset NHS Foundation Trust, Somerset

Stephen Murphy, Imperial College Healthcare NHS Trust, London

Harri Sivasathiaseelan, Homerton University Hospital NHS Foundation Trust, London

Richard Sylvester, Homerton University Hospital NHS Foundation Trust, London

Abdul Al-Saleh, The Royal Wolverhampton NHS Trust, Wolverhampton

Rhys Roberts, Addenbrookes Hospital, Cambridge University Hospitals NHS Foundation Trust, Cambridge

Kannan Nithi, Northampton General Hospital, Northampton General Hospital NHS Trust, Northampton

Lahiru Handdunnethi, John Radcliffe Hospital, Oxford University Hospitals NHS Foundation Trust, Oxford

Kate Wannop, Kings College Hospital NHS Foundation Trust, London

Amit Batla, Royal Free Hospital NHS Foundation Trust, London

Anna Sadnicka, Royal Free Hospital NHS Foundation Trust, London

Jananee Sivaganasundaram, Epsom and St Helier University Hospitals NHS Trust, Epsom

Tatyana Yermakova, Leeds General Infirmary, Leeds Teaching Hospitals NHS Trust, Leeds

Ravi Dasari, Kent and Canterbury Hospital, East Kent Hospitals University NHS Foundation Trust

Graziella Quattrocchi, North Middlesex Uniersity Hospital NHS Trust, London

Harriet Ball, North Bristol NHS Trust, Bristol

Rebecca Cooper, Princess Royal Hospital, Haywards Heath, University Hospitals Sussex NHS Foundation Trust

Daniel Whittam, Manchester Centre for Clinical Neurosciences, Salford Royal NHS Foundation Trust, Manchester

Mohanned Mustafa, Manchester Centre for Clinical Neurosciences, Salford Royal NHS Foundation Trust, Manchester

Gabriel Yiin, Great Western Hospitals NHS Foundation Trust, Swindon

Shayan Ashjaei, Kings College Hospital NHS Foundation Trust, London

Andrew J. Westwood, Tunbridge Wells Hospital, Maidstone and Tunbridge Wells NHS Trust, Kent

Michelle Dsouza, Kent and Canterbury Hospital, East Kent Hospitals University NHS Foundation Trust

Eng Chuan Foo, Calderdale Royal Hospital, Calderdale and Huddersfield NHS Foundation Trust, Halifax

Shwe Zin Tun, Calderdale Royal Hospital, Calderdale and Huddersfield NHS Foundation Trust, Halifax

Khine Khine Lwin, Calderdale Royal Hospital, Calderdale and Huddersfield NHS Foundation Trust, Halifax

Gorande Kanabar, East & North Hertfordshire NHS Trust

